# Waterlogging exposure and mental well-being: a cross-sectional study in rural southwest Bangladesh

**DOI:** 10.64898/2026.08.04.26359716

**Authors:** Lucie Clech, Emmanuel Bonnet, DM Rezoan, Mollah M Kabir, Mary Shenk, Valéry Ridde

## Abstract

**Background:** Waterlogging, a form of chronic, stagnant flooding, is increasing in the Ganges-Brahmaputra delta in Bangladesh due to the compounding effects of land-use change, including the expansion of brackish shrimp farming, poor water management, and changing rainfall regimes. Its effects are negatively impacting livelihoods and health; its association with mental wellbeing is less known.

**Methods:** We hypothesised that 1-recent waterlogging, 2-social disadvantage (women, older individuals, the poorest, the least educated, those with chronic illness, and religious minorities) would be associated with lower wellbeing, and 3-chronic illness would modify the association between waterlogging and mental wellbeing. 1260 respondents from 595 households in Tala upazila, southwest Bangladesh, were interviewed about their mental wellbeing, chronic illness, and exposure to waterlogging in the 12 months prior to data collection, in August and September 2022. Associations between WHO-5 wellbeing scores and waterlogging and covariates were assessed using multi-level linear mixed-effects models with household random effects and cluster fixed effects.

**Results:** Our results confirm our hypotheses: wellbeing was lower among disadvantaged groups and chronic health vulnerability modifies the association between waterlogging and wellbeing: waterlogging exposure was associated with 18.31-point lower WHO-5 scores among individuals with chronic illness (95% CI -26.45 to -10.17), an association markedly attenuated among those without chronic illness (interaction β=14.21, 95% CI 5.48 to 22.95, p=0.001).

**Conclusion:** These results suggest that chronic illness may increase vulnerability to the mental health burden associated with waterlogging. As waterlogging is increasing, policies addressing both its environmental drivers and the needs of vulnerable populations should be considered.

**Competing interests:** None declared

This study is funded by the French National Research Agency (ANR) as part of the French presidential call “Make Our Planet Great Again” (MOPGA), ANR-18-MPGA-0010. The funders had no role in study design, data collection and analysis, publication decision, or manuscript preparation. The Grant recipient was VR.

## 1. Introduction

Flooding is among the most frequent natural disasters globally (1), yet its consequences for subjective wellbeing and mental health remain poorly characterised compared with physical health outcomes in rural populations of low- and middle-income countries (2–4). Bangladesh is among the most climate-vulnerable countries in the world (5), with the southwest delta particularly exposed to waterlogging, a form of stagnant flooding, resulting from decades of attempts to control a naturally dynamic deltaic ecosystem through land and water engineering, including mangrove clearance, polders, dams, river embankments and the expansion of brackish shrimp farming (6–8). Waterlogging inundates land and properties, disrupts livelihoods and constrains access to healthcare, leaving communities exposed to its consequences well beyond the flood season (6,7,9,10).

Poorer mental health outcomes are associated with chronic illness (11,12), age, education, and wealth in rural Bangladesh (13) and with religious minority status among Bangladeshi women (14). In North East Bangladesh, flooding has been associated with depression among rural women, persisting up to 2.5 years after the event and operating through pathways of food insecurity and household indebtedness (15). The impact of environmental change on populations is not uniform but rather conditioned by pre-existing inequalities (16,17). Low socioeconomic status, limited education and physical health problems are associated with poor mental health outcomes after exposure to floods (4). In coastal Bangladesh, environmental stressors linked to sea-level rise have been associated with increased psychological distress, depression, anxiety and stress, with older adults, women, and individuals from lower income in the most exposed communities disproportionately affected (18,19).

Social position shapes not only exposure to environmental stressors but also the resources and capacity to cope with their consequences. Socio-economic resources, including income, education, social connections, determine who is protected from environmental harm (16,20) while the embodiment of social disadvantage over the life course shapes the biological susceptibility with which individuals face additional stress (21).

Subjective wellbeing remains underexplored in the context of environmental change. Yet, environmental stressors may affect wellbeing across the full spectrum, including sub-clinical states below diagnostic thresholds. No quantitative study has examined the association between subjective wellbeing, waterlogging exposure, and whether this association differs by socio-economic status and health.

Building on two prior studies from the same region (7,9), we hypothesised that (H1) waterlogging exposure would be negatively associated with WHO-5 Well-Being Index (22) in Tala Upazila, southwest Bangladesh; (H2) wellbeing would be lower among women, older individuals, the poorest, the least educated, those with chronic illness, and religious minorities, reflecting their social position, autonomy, and their comparatively lower access to social, economic and health resources (16,20); and (H3) the waterlogging-wellbeing association would be more pronounced among those with chronic illness, who may face compounded disadvantage when exposed to additional environmental stress (16,20,21). Interactions with gender, age, education and wealth were also explored.

## 2. Methods

### Ethics approval and consent to participate

The Institutional Review Board (IRB) of the BRAC James P. Grant School of Public Health, BRAC University granted ethics approval (ref: IRB-19 November’20–050) in Bangladesh. Study information was provided to the respondents prior to data collection, and written informed consent was sought before each interview.

### Data source and study settings

This study is part of the ClimHB project, which explores the links between climate change, migration, and health system resilience (23,24). The data come from a cross-sectional household survey in Tala Upazila, Bangladesh, collected in August and September 2022. Tala Upazila, a 344.15 km^2^ administrative unit, is located in the Satkhira subdistrict, a rural territory prone to waterlogging in the Ganges-Brahmaputra delta (7,8).

### Data

We interviewed a total of 1268 adult individuals from 596 households. Only questionnaires with full information were included (N=1260 respondents, 595 households). Sampling is presented in SM1

### Measures

Descriptive statistics are presented in Supplementary Materials 1 (SM 1).

### Exposure variable

Waterlogging exposure was defined as household-reported experience of at least one waterlogging event in the preceding 12-month recall period to balance recall accuracy and coverage of the current and last rainy season, given that data collection occurred during the Monsoon season but a drought year. Flood self-reporting is more accurate than satellite-based evaluation (25), and recall accuracy remains high even after longer recall periods (26,27).

### Outcome variables

Subjective mental wellbeing was measured using the WHO-5 mental wellbeing index, a five-item questionnaire (22) validated in Bangladesh (28). The aggregated score ranged from 0 to 100, where a lower score represents worse mental well-being. The use of a continuous score allows for a more nuanced measure of variation in mental wellbeing in the context of environmental change.

### Additional study variables

Covariate coefficients (H2) reflect group differences from the same adjusted model used for H1, not separate causal effects for each variable. An individual was classified as having reported a chronic illness if they had reported any chronic condition lasting more than three consecutive months (9). Chronic health is known to be associated with worse mental health outcomes (11,12). Acute symptoms (<1 month) were not included, given their diverse aetiologies and severities. Other individual and household-level characteristics were used as covariates, both to control for confounding and to examine their association with the outcome of interest. Age was dichotomised at 50 years to distinguish younger from older individuals (9). Household economic status was captured by taking the log of household income to account for diminishing marginal returns in wellbeing (29). Alternative models with other wealth specifications are presented and discussed in Supplementary Materials (SM) 2. Individual microcredit use was included as a binary covariate. Gender (male/female), education (none/primary/secondary/higher), head-of-household status (yes/no) and religion (Muslim/Hindu) were also included. Household size was entered as a continuous variable to limit model complexity, given the data’s multilevel structure and the number of categorical predictors already included.

Given the limited number of exposed individuals (n=71), interaction terms were tested individually for gender, age, wealth, education and chronic health status rather than simultaneously, and only statistically significant interactions were retained in the final model.

Trained public health interviewers used digital data collection tools that automatically checked responses for basic consistency and plausibility. Data were collected by a team of 20 trained field assistants using SurveyCTO software, version 2.70.

### Reference coding

For waterlogging, the reference was no exposure. The reference categories were assigned to the most vulnerable groups within each variable (women, older individuals, the poorest, less educated, with chronic illness, religious minority, not the head of household). Coefficients represent the relative advantage of not being vulnerable, shifting the interpretative frame from individual risk (30) to relative structural inequality (31). For individual credit use, the reference was selected as a non-user.

### Statistical analysis

Descriptive statistics are presented in (SM1) at the individual level, with additional information on cluster-level exposure in (SM 3). Associations between waterlogging exposure and WHO-5 wellbeing scores were assessed using multi-level linear mixed-effects models with household random effects and cluster fixed effects, using restricted maximum likelihood (REML) in R (lme4 package). A three-level data structure was considered: individuals nested within households; households nested within clusters. The rationale for cluster fixed effects versus cluster random effects is presented in SM 4.

As a sensitivity analysis, generalised linear mixed logistic regression models were estimated using the same covariates at the validated clinical thresholds of WHO-5 ≤50 (poor well-being warranting further assessment for depressive symptoms) and ≤28 (severely poor, major depression screening criteria) (22). The models (Table 2) were fitted using maximum likelihood. To facilitate convergence, models were fitted using the Laplace approximation (nAGQ=0) and the bobyqa optimizer with increased iteration limits to ensure stable model convergence. Risk ratios were approximated from odds ratios using the method of Zhang and Yu (32).

**Table 2.** Multilevel linear mixed-effects models, WHO-5 wellbeing index score.

|  | Model 1 |  | Model of interest:<br>M1+interaction |  | Model 2 |  | Model 3 |  |
| --- | --- | --- | --- | --- | --- | --- | --- | --- |
| Sample | Full sample |  | Full sample |  | Clusters with<br>waterlogging |  | Clusters without<br>waterlogging |  |
| Variable (reference category) | $\beta$_M1[IC 95%] | p_M1 | $\beta$_M1i[IC 95%] | p_M1i | $\beta$_M2[IC 95%] | p_M2 | $\beta$_M3[IC 95%] | p_M3 |
| (Intercept) | -20.21 [-42.69, 2.26] | 0.078 | -18.34 [-40.68, 3.99] | 0.107 | 4.67 [-31.37, 40.72] | 0.798 | -26.75 [-55.34, 1.85] | 0.067 |
| Exposure to waterlogging: exposed (not) | -8.95 [-14.77, -3.14] | 0.003 | -18.31 [-26.45, -10.17] | <0.001 | -17.25 [-26.19, -8.32] | <0.001 |  |  |
| Age group: 18–50 (51+) | 5.1 [2.61, 7.59] | <0.001 | 5.02 [2.54, 7.51] | <0.001 | 5.67 [1.2, 10.14] | 0.013 | 4.97 [1.95, 7.99] | 0.001 |
| Gender: male (female) | 3.85 [0.88, 6.83] | 0.011 | 3.95 [0.98, 6.91] | 0.009 | 3.14 [-3.02, 9.29] | 0.316 | 4.1 [0.65, 7.56] | 0.020 |
| Religion: Muslim (Hindu) | 4.58 [1.39, 7.77] | 0.005 | 4.77 [1.6, 7.93] | 0.003 | 1.72 [-4.97, 8.4] | 0.613 | 5.68 [2.04, 9.32] | 0.002 |
| Education: primary (No education) | 5.2 [2.2, 8.2] | <0.001 | 5.11 [2.12, 8.1] | <0.001 | 4.46 [-0.7, 9.62] | 0.090 | 5.19 [1.5, 8.87] | 0.006 |
| Education: secondary (No education) | 7.81 [4.82, 10.81] | <0.001 | 7.7 [4.72, 10.69] | <0.001 | 7.9 [2.74, 13.06] | 0.003 | 7.34 [3.65, 11.03] | <0.001 |
| Education: higher (No education) | 8.88 [2.99, 14.76] | 0.003 | 9.08 [3.21, 14.95] | 0.002 | 6.45 [-4.47, 17.37] | 0.246 | 9.49 [2.45, 16.53] | 0.008 |
| Household monthly income (log) | 5.38 [2.96, 7.81] | <0.001 | 5.24 [2.83, 7.65] | <0.001 | 3.79 [-0.17, 7.74] | 0.060 | 6.15 [3.04, 9.26] | <0.001 |
| Household size: number of individuals | -0.86 [-1.85, 0.13] | 0.089 | -0.84 [-1.82, 0.14] | 0.093 | -0.86 [-2.76, 1.04] | 0.375 | -0.89 [-2.06, 0.27] | 0.133 |
| Microcredit: user (not user) | -1.31 [-3.42, 0.8] | 0.225 | -1.23 [-3.33, 0.87] | 0.252 | 0.71 [-3.07, 4.49] | 0.711 | -1.92 [-4.48, 0.64] | 0.141 |
| Household headship: head (not head) | 0.61 [-2.41, 3.63] | 0.693 | 0.51 [-2.51, 3.52] | 0.742 | 2.74 [-3.56, 9.05] | 0.392 | -0.19 [-3.69, 3.32] | 0.917 |
| Chronic illness status: no chronic (chronic illness) | 9.92 [7.76, 12.09] | <0.001 | 9.09 [6.87, 11.31] | <0.001 | 8.72 [3.72, 13.72] | <0.001 | 9.2 [6.67, 11.72] | <0.001 |
| Waterlogging × No chronic illness | | | 14.21 [5.48, 22.95] | 0.001 | 13.45 [3.88, 23.01] | 0.006 | | |
| Cluster 2 | 9.41 [3.89, 14.92] | <0.001 | 9.17 [3.69, 14.65] | 0.001 |  |  |  |  |
| Cluster 3 | 0.74 [-4.59, 6.08] | 0.784 | 0.69 [-4.6, 5.99] | 0.797 |  |  | 0.75 [-4.59, 6.09] | 0.782 |
| Cluster 4 | -0.81 [-6.2, 4.57] | 0.768 | -0.94 [-6.28, 4.4] | 0.730 |  |  | -0.91 [-6.32, 4.5] | 0.741 |
| Cluster 5 | -5.01 [-10.55, 0.54] | 0.077 | -5.02 [-10.52, 0.48] | 0.073 | -13.02 [-19.59, -6.46] | <0.001 |  |  |
| Sample | Full sample |  | Full sample |  | Clusters with<br>waterlogging |  | Clusters without<br>waterlogging |  |
| Variable (reference category) | $\beta$_M1[IC 95%] | p_M1 | $\beta$_M1i[IC 95%] | p_M1i | $\beta$_M2[IC 95%] | p_M2 | $\beta$_M3[IC 95%] | p_M3 |
| Cluster 6 | -6.24 [-11.74, -0.75] | 0.026 | -6.42 [-11.87, -0.96] | 0.021 |  |  | -6.39 [-11.96, -0.83] | 0.024 |
| Cluster 7 | 2.83 [-2.69, 8.34] | 0.314 | 2.73 [-2.74, 8.2] | 0.328 |  |  | 2.48 [-3.08, 8.04] | 0.381 |
| Cluster 8 | 0.87 [-4.43, 6.16] | 0.748 | 0.92 [-4.33, 6.17] | 0.731 |  |  | 1.29 [-4, 6.58] | 0.632 |
| Cluster 9 | 10.21 [4.39, 16.03] | <0.001 | 10.55 [4.78, 16.33] | <0.001 | 1.86 [-4.46, 8.17] | 0.562 |  |  |
| Cluster 10 | 0.01 [-5.32, 5.35] | 0.996 | -0.08 [-5.37, 5.22] | 0.977 |  |  | -0.14 [-5.48, 5.2] | 0.958 |
| *Number of observations* | 1260 |  | 1260 |  | 369 |  | 891 |  |
| *Number of households* | 595 |  | 595 |  | 177 |  | 418 |  |
| *Residual variance* | 218.51 |  | 218.51 |  | 188.61 |  | 231.09 |  |
| *ICC (household level)* | 0.33 |  | 0.323 |  | 0.394 |  | 0.3 |  |
| *Marginal R<sup>2</sup>* | 0.202 |  | 0.208 |  | 0.279 |  | 0.171 |  |
| *Conditional R<sup>2</sup>* | 0.465 |  | 0.464 |  | 0.563 |  | 0.419 |  |
| *AIC* | 10749 |  | 10736.1 |  | 3094.1 |  | 7618.1 |  |

Notes: Models estimated using multilevel linear mixed-effects model regression Reference categories: no waterlogging exposure, age $\geq 51$ years, female, Hindu religion, no formal education, not microcredit user, not head of household, having a chronic illness.
Cluster 1 as reference for cluster fixed effects.
Alternative models to the model of interest are presented for comparison in Supplementary Materials 2. Results are robust across the four alternative models (a-model of interest (M1 + interaction), b-model with quadratic terms, c-model with wealth index quartiles, and d-model with wealth index score).
M2 on two clusters (2,9) is presented in Supplementary Materials 5: results are robust across M2 models.
Non-significant interactions are presented in Supplementary Materials 6.

**Table 3:** Generalised linear mixed logistic regression models. WHO-5 well-being index

| | WHO-5 $\leq 50$ (with interaction) | | Model of interest<br>WHO-5 $\leq 50$ (no interaction) | | WHO-5 $\leq 28$ (with interaction) | | Model of interest<br>WHO-5 $\leq 28$ (no interaction) | |
| --- | --- | --- | --- | --- | --- | --- | --- | --- |
| Variable (reference category) | OR [95% CI] | p-value | OR [95% CI] | p-value | OR [95% CI] | p-value | OR [95% CI] | p-value |
| Exposure to waterlogging: exposed (not) | 4.01 [1.13, 14.19] | 0.031 | 2 [0.92, 4.33] | 0.079 | 8.09 [2.45, 26.74] | <0.001 | 5.02 [1.96, 12.87] | <0.001 |
| Age group: 18–50 (51+) | 0.61 [0.42, 0.88] | 0.008 | 0.6 [0.42, 0.87] | 0.007 | 0.51 [0.34, 0.77] | 0.001 | 0.51 [0.34, 0.77] | 0.001 |
| Gender: male (female) | 0.65 [0.42, 1.02] | 0.060 | 0.66 [0.42, 1.03] | 0.066 | 0.74 [0.44, 1.25] | 0.261 | 0.75 [0.44, 1.27] | 0.282 |
| Religion: Muslim (Hindu) | 0.45 [0.3, 0.69] | <0.001 | 0.46 [0.3, 0.7] | <0.001 | 0.72 [0.44, 1.18] | 0.192 | 0.73 [0.45, 1.2] | 0.215 |
| Education: primary (No education) | 0.62 [0.4, 0.97] | 0.036 | 0.62 [0.4, 0.97] | 0.035 | 0.65 [0.4, 1.05] | 0.075 | 0.65 [0.4, 1.04] | 0.073 |
| Education: secondary (No education) | 0.4 [0.26, 0.62] | <0.001 | 0.4 [0.26, 0.62] | <0.001 | 0.45 [0.28, 0.74] | 0.002 | 0.45 [0.28, 0.74] | 0.002 |
| Education: higher (No education) | 0.52 [0.22, 1.21] | 0.128 | 0.53 [0.23, 1.23] | 0.137 | 0.6 [0.21, 1.69] | 0.332 | 0.61 [0.22, 1.72] | 0.354 |
| Household monthly income (log) | 0.51 [0.37, 0.72] | <0.001 | 0.51 [0.36, 0.71] | <0.001 | 0.58 [0.39, 0.87] | 0.008 | 0.57 [0.39, 0.86] | 0.006 |
| Household size: number of individuals | 1.12 [0.98, 1.28] | 0.088 | 1.12 [0.98, 1.28] | 0.084 | 1.13 [0.98, 1.31] | 0.103 | 1.14 [0.98, 1.32] | 0.095 |
| Microcredit: user (not user) | 1.34 [1, 1.81] | 0.052 | 1.35 [1, 1.82] | 0.048 | 0.97 [0.67, 1.39] | 0.852 | 0.97 [0.67, 1.4] | 0.879 |
| Household headship: head (not head) | 0.92 [0.58, 1.44] | 0.707 | 0.91 [0.58, 1.43] | 0.677 | 0.74 [0.43, 1.26] | 0.265 | 0.73 [0.43, 1.24] | 0.245 |
| Chronic illness status: no chronic (chronic illness) | 0.4 [0.29, 0.55] | <0.001 | 0.37 [0.27, 0.52] | <0.001 | 0.36 [0.25, 0.51] | <0.001 | 0.34 [0.24, 0.48] | <0.001 |
| Waterlogging × No chronic illness | 0.36 [0.09, 1.47] | 0.156 | | | 0.4 [0.1, 1.65] | 0.203 | | |
| Cluster 2 | 0.37 [0.18, 0.77] | 0.008 | 0.36 [0.17, 0.76] | 0.007 | 0.29 [0.11, 0.79] | 0.016 | 0.29 [0.11, 0.77] | 0.013 |
| Cluster 3 | 0.94 [0.47, 1.89] | 0.862 | 0.94 [0.46, 1.89] | 0.855 | 0.71 [0.31, 1.63] | 0.420 | 0.7 [0.31, 1.62] | 0.409 |
| Cluster 4 | 1.02 [0.51, 2.06] | 0.957 | 1.01 [0.5, 2.05] | 0.977 | 1.3 [0.59, 2.85] | 0.520 | 1.28 [0.58, 2.82] | 0.540 |
| Cluster 5 | 1.75 [0.85, 3.63] | 0.132 | 1.75 [0.84, 3.64] | 0.134 | 1.5 [0.66, 3.42] | 0.330 | 1.49 [0.66, 3.4] | 0.338 |

| Variable (reference category) | WHO-5 $\leq 50$ (with interaction) | | Model of interest<br>WHO-5 $\leq 50$ (no interaction) | | WHO-5 $\leq 28$ (with interaction) | | Model of interest<br>WHO-5 $\leq 28$ (no interaction) | |
| --- | --- | --- | --- | --- | --- | --- | --- | --- |
|  | OR [95% CI] | p-value | OR [95% CI] | p-value | OR [95% CI] | p-value | OR [95% CI] | p-value |
| Cluster 6 | 2.26 [1.08, 4.73] | 0.031 | 2.25 [1.07, 4.73] | 0.033 | 1.25 [0.55, 2.84] | 0.587 | 1.23 [0.54, 2.79] | 0.617 |
| Cluster 7 | 0.66 [0.32, 1.35] | 0.255 | 0.65 [0.32, 1.35] | 0.248 | 1.42 [0.63, 3.24] | 0.401 | 1.41 [0.62, 3.21] | 0.413 |
| Cluster 8 | 0.8 [0.4, 1.6] | 0.533 | 0.81 [0.4, 1.61] | 0.543 | 0.87 [0.39, 1.91] | 0.724 | 0.87 [0.39, 1.92] | 0.727 |
| Cluster 9 | 0.34 [0.15, 0.73] | 0.006 | 0.34 [0.16, 0.74] | 0.006 | 0.21 [0.07, 0.62] | 0.004 | 0.22 [0.08, 0.63] | 0.005 |
| Cluster 10 | 0.99 [0.49, 1.97] | 0.966 | 0.98 [0.49, 1.96] | 0.948 | 0.82 [0.36, 1.84] | 0.625 | 0.81 [0.36, 1.82] | 0.606 |
| *Number of observations* | *1260* |  | *1260* |  | *1260* | *1260* | *1260* |  |
| *Number of households* | *595* |  | *595* |  | *595* | *595* | *595* |  |
| *Household-level variance* | *1.18* |  | *1.21* |  | *0.97* | *0.97* | *0.97* |  |
| *AIC* | *1542.3* |  | *1542.9* |  | *1082.6* | *1082.6* | *1082.6* |  |

OR: Odds Ratio; CI: 95% Confidence Interval. OR < 1 indicates lower odds of poor wellbeing relative to the reference category, whereas OR > 1 indicates higher odds of poor wellbeing.
WHO-5 $\leq 50$ : validated screening threshold for poor well-being/depression symptoms
WHO-5 $\leq 28$ : threshold for severe poor well-being.
Interaction terms chronic X waterlogging are not statistically significant for both WHO-5 threshold, and thus not added in the models of interest
Models estimated using generalised linear mixed-effects logistic regression
(glmer, bobyqa optimiser, lme4 package, R version 4.3.0).
Reference categories: no waterlogging exposure, age $\geq 51$ years, female, Hindu religion, no formal education, not microcredit user, not head of household, having a chronic illness.
Cluster 1 as reference for cluster fixed effects.

### Spatial analysis

To assess spatial heterogeneity, we calculated global Moran’s I to test for overall spatial autocorrelation in wellbeing scores. Local Indicators of Spatial Association (LISA) were then used to identify significant clusters and spatial outliers. These analyses were applied separately to observed wellbeing scores, model-predicted values, and residuals. Statistical significance was assessed using permutation tests.

Statistical significance was assessed at α = 0.05, for all models.

We used the STROBE reporting guideline when drafting and editing the manuscript.

## 3. Results

### 3.1 Association between waterlogging and well-being

A statistically significant negative association between reported waterlogging exposure in the past 12 months and well-being (WHO-5 index) is robust across all models (Tables 1 and 2 and SM 2, 4, 5, 6).

In unadjusted analyses (SM1), the mean WHO-5 score in the sample was 48.5 (SD 20.1), 52.3% of individuals scored≤50, and 18.8% scored ≤28. The well-being index did not differ significantly between waterlogging-exposed (mean 46.8, SD 25.5) versus non-exposed individuals (mean 48.6, SD 19.8) (p=0.559, p=0.108, and p=0.689). However, after adjustment, exposure was associated with reduced well-being (Table 1), poor well-being, and severely poor wellbeing (Table 2).

In two-level linear mixed-effects models, exposure was associated with a statistically significant 9-point reduction in individuals’ wellbeing scores (β = −8.95, 95%CI: -14.77 to - 3.14, p = 0.003), Model 1, Table 1). Our model of interest is the model with the interaction term waterlogging × chronic illness (Model M1+ interactions; Table 1). In this model, exposure was associated with an18.3-point reduction in wellbeing compared to no exposure (β = −18.31, 95% CI: −26.45, −10.17, p < 0.001). The results were robust across alternative models (SM 2), for the models with 1-quadratic income terms, 2-wealth index quartile and 3-wealth index score.

In Model M2 (Table 1), performed only on the three clusters with declared waterlogging, among individuals with chronic illness (reference group), exposure was associated with a 17.3-point decrease in wellbeing compared to no exposure (β = −17.25, 95% CI: −26.19 to −8.32, p < 0.001, n=369 respondents). Results were also robust in an alternative model restricted to two clusters (clusters 2 and 9) only: (β = −18.1, 95% CI: −28.29 to −7.91, p < 0.001, n=250 respondents; SM 5).

When specifically looking at poor wellbeing (WHO-5 index ≤ 50), a generalised linear mixed logistic regression model shows that waterlogging is (nearly significant, p=0.079, Table 2) associated with 2 times the odds of poor wellbeing and for WHO-5 index ≤ 28 at 5.02 times greater odds of reporting severely poor wellbeing when their households were exposed (WHO-5 ≤28; OR = 5.02, 95% CI [1.96, 12.87], p < 0.001), corresponding to an estimated risk ratio of 1.615 (95% CI [1.304, 1.780]). Exposed individuals were approximately 62% more likely to report poor well-being compared to non-exposed individuals after adjustment. These results are robust across models; in alternative generalised linear mixed logistic regression models using quadratic terms for standardised income, the associations between waterlogging and wellbeing are even stronger and all statistically significant (WHO-5 ≤50; OR = 2.43, 95% CI [1.03, 5.69], p < 0.042; WHO-5 ≤28; OR = 7.25, 95% CI [2.31, 22.79], p < 0.001, see SM 7).

The linear mixed-effects models, performed on the full continuous WHO-5 score (Table 1), suggest a more pronounced association, as among those with chronic illness waterlogging-exposed individuals scored, on average, 18.31 points lower on the WHO-5 (β = −18.31, 95% CI: −26.45, −10.17, p <0.001) which cannot be fully captured by the logistic regression (Table 2). The logistic model is used here for sensitivity analysis and should be interpreted as a conservative estimate of the association between waterlogging and poor well-being. The following results are presented only for the multilevel linear models (Table 1 and SM 2, 5).

### 3.2 Mental well-being varies across socio-demographic and economic groups

Higher wellbeing scores were observed among younger individuals (aged 18-50) compared to elders (aged 51 years and above), (β ≈ 4.9-5.7, p ≤ 0.01 in all models, Table 1) as well as among men compared to women (β ≈ 3.8 to 4.1, p ≤ 0.05 for all models except M2, NS, Table 1) and Muslims compared to members of the Hindu minority (β ≈4.5 to 5.7, p ≤ 0.005 for all models except M2, NS, Table 1). Translating those results into wellbeing scores, this means that younger individuals reported about 5 to 6 more points in wellbeing scores compared to older individuals, men reported about 4 points more than women, and Muslims about 4 to 6 points more than Hindus.

Individuals with primary, secondary or higher levels of education had significantly higher well-being scores compared to those with no formal education (Primary/no education: β ≈ 5.1 to 5.2, p < 0.006 for all models except M2, NS; Secondary/no education: β ≈ 7.3 to 7.9, p ≤ 0.003 for all four models; Higher/no education: β ≈ 8.8 to 9.5, p ≤ 0.008 for all models except M2, NS, Table 1).

Higher household income (log-transformed monthly income) was associated with improved wellbeing score (β ≈ 5.2 to 6.2, p < 0.001 for all models except M2, near-significant at p = 0.060, Table 1). Alternative wealth variables were tested (SM2) but did not improve the model. These results suggest that socio-economic advantage is associated with higher well-being, whereas more vulnerable or minority groups tend to experience lower well-being. In contrast, household size, individual access to micro-credit and household headship were not significantly associated with wellbeing (Table 1).

Gender, religion, household income were no longer statistically significant in model M2 (reduced sample of exposed clusters only), suggesting reduced statistical power rather than differences in associations (Table 1). M2 was also performed on clusters 2 and 9 (cluster 5 being removed) and results were robust (SM 5).

### 3.3 Mental well-being varies with chronic illness status

Absence of chronic illness in the past 2 months is associated with higher wellbeing scores compared to individuals reporting chronic illness for the four models (β ≈8.7 to 9.9, p < 0.001, Table 1). In other words, chronically ill individuals tend to experience lower well-being, as measured by the WHO-5 score. In Model M1+interactions, among individuals with no exposure (reference group), not reporting chronic illness was associated with a 9.1-point increase in wellbeing compared to reporting chronic illness (β = 9.09, 95% CI: 6.87, 11.31, p < 0.001, Table 1). In Model M2, among individuals with no exposure (reference group), not reporting chronic illness was associated with a 8.7-point increase in wellbeing compared to reporting chronic illness (β = 8.72, 95% CI: 3.72 to 13.72, p < 0.001, Table 1).

### 3.4 The negative association of waterlogging and wellbeing is stronger among vulnerable groups

The decrease in wellbeing associated with waterlogging is modest among those without chronic illness, but stronger among those with chronic illness (Figure 1). The negative association with waterlogging was attenuated by 14.21 points (β = 14.21, 95% CI: 5.48, 22.95, p < 0.001, Table 1) among participants without chronic illness, indicating that chronic illness amplifies the negative association of waterlogging and wellbeing. Interaction terms between waterlogging and other covariates were not significant and were not included in the model. Alternative models with interaction terms for age group, gender, income, and education are presented in SM 6.

**Figure 1:**
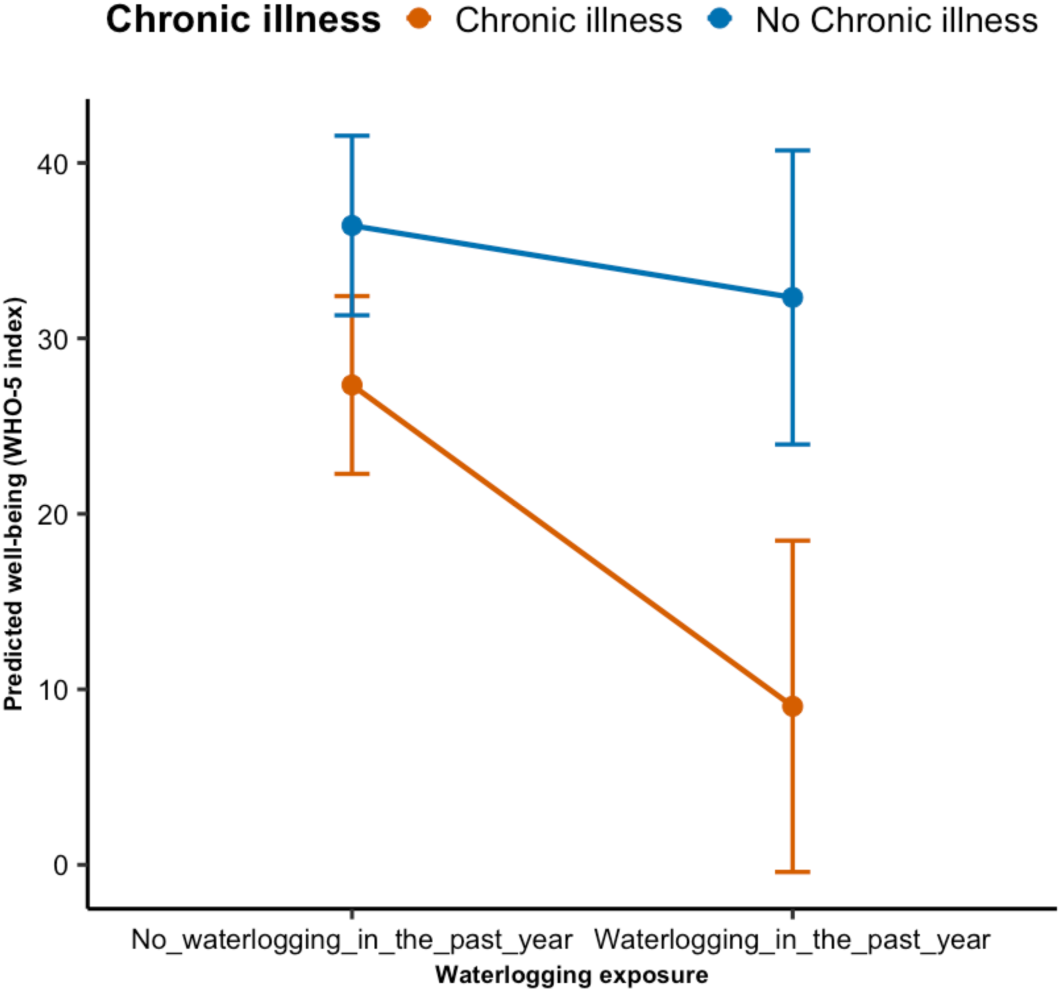
Predicted wellbeing scores by waterlogging exposure and chronic illness status (interaction model) <u>Notes:</u> Predicted wellbeing scores (WHO-5 index) derived from mixed-effects regression models (model M1 with interaction, Table 1), showing the interaction effect of waterlogging exposure and chronic illness status. Error bars represent 95%CI. The predicted values are below the clinical WHO-5 index ≤ 50 threshold.

### 3.5 Spatial heterogeneity

Observed WHO-5 wellbeing showed weak spatial autocorrelation (Moran’s I=0.075), with a near-random distribution. Model-predicted values present stronger spatial autocorrelation (I=0.269), with High-High and Low-Low groupings (Figure 2). As predictions incorporate both individual, household and cluster characteristics, this suggests that the spatial heterogeneity in wellbeing reflects both household profiles and unmeasured cluster-level factors, invisible in raw scores. Residuals showed almost no spatial autocorrelation (I=0.015), indicating that our model adequately captures the spatial structure in wellbeing.

**Figure 2:**
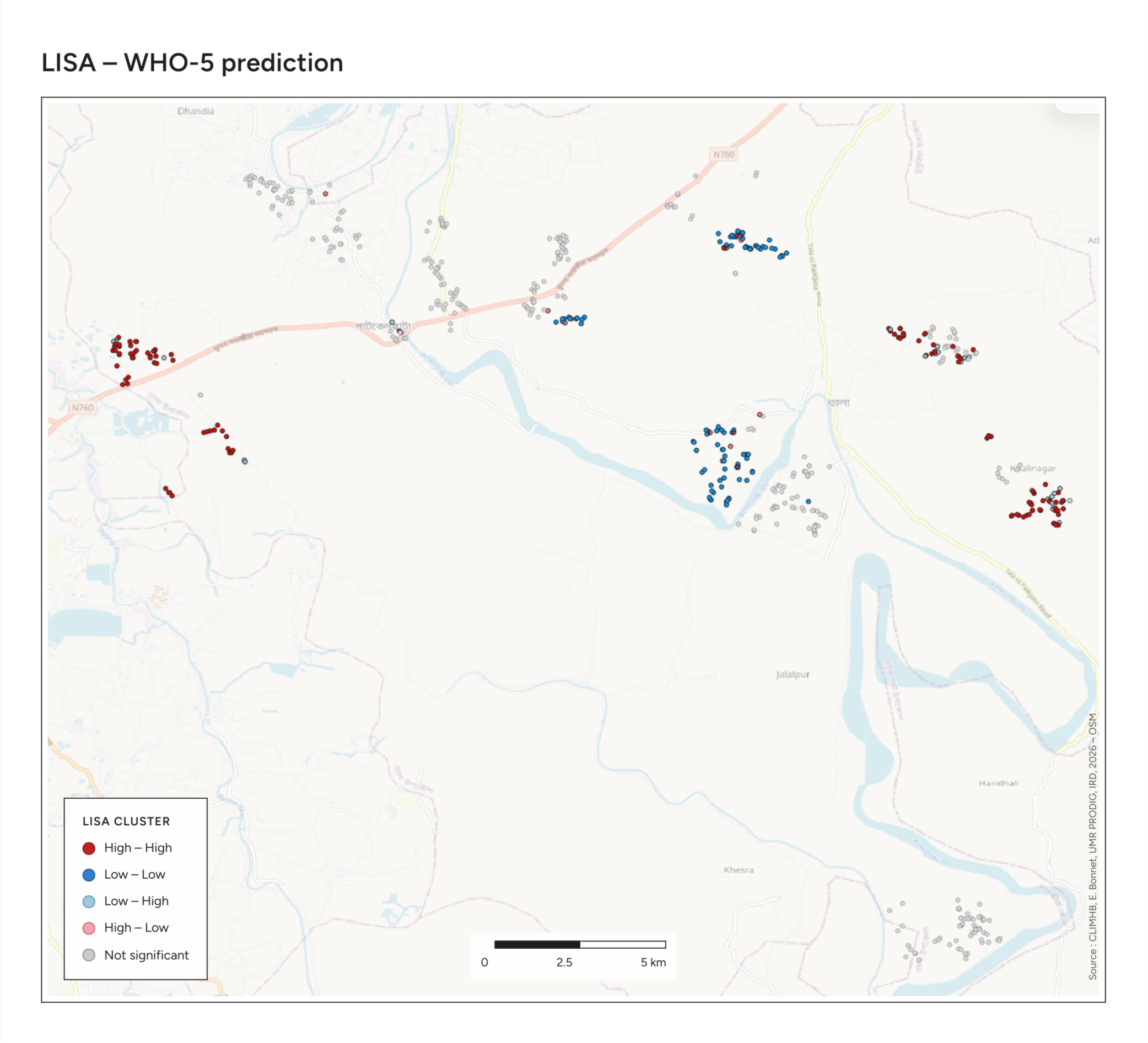
Local spatial clustering (LISA) of predicted WHO-5 wellbeing scores in Tala Upazila, Bangladesh Notes: Local Indicators of Spatial Association (LISA) were applied to model-predicted WHO-5 wellbeing scores. Red (High-High) and blue (Low-Low) clusters indicate significant spatial clustering of higher and lower predicted wellbeing (respectively). Light red (High-Low) and light blue (Low-High) indicate spatial outliers. Grey points are not statistically significant. OSM.

## Discussion

Our hypotheses are confirmed. Waterlogging exposure (H1) was associated with lower wellbeing (β=-8.95, 95% CI -14.77 to -3.14, p=0.003). Consistent with H2, an advantage in socio-demographic and economic status is associated with better wellbeing. Men had 3.85 points higher wellbeing than women (95% CI 0.88 to 6.83, p=0.011); individuals aged 18-50 had 5.10 points higher wellbeing than those aged 51 and above (95% CI 2.61 to 7.59, p<0.001); Muslims had 4.58 points higher wellbeing than Hindus (95% CI 1.39 to 7.77, p=0.005); those with primary, secondary or higher education had 5.20 to 8.88 points higher wellbeing than those with no education (all p<0.01); each unit increase in log household income was associated with a 5.38-point increase in wellbeing (95% CI 2.96 to 7.81, p<0.001); and individuals without chronic illness had 9.92 point higher wellbeing than those with chronic illness (95% CI 7.76 to 12.09, p<0.001). These group differences are adjusted for the same model used to estimate the waterlogging association, and are not independent causal estimates. Consistent with H3, the negative association between waterlogging and wellbeing was amplified among individuals with chronic illness, with an association 14.21 points more pronounced than among those without (95% CI 5.48 to 22.95, p=0.001).

Although the WHO-5 scale has no validated clinical interpretation of point difference, only clinically validated thresholds at WHO-5 ≤50 and at WHO-5 ≤ 28 for poor wellbeing and severely poor wellbeing, in a population characterised by poor wellbeing (mean WHO-5 48.5 (SD 20.1)), even a small but statistically significant difference in wellbeing can matter at the population level.

Results about waterlogging exposure are consistent with the mental health literature on flooding (3). Our findings further extend this literature by focusing on wellbeing rather than clinical outcomes, and on waterlogging, a type of flooding that occurs repeatedly and often for extended periods rather than sporadically. Although our cross-sectional design captures at least one occurrence in the past 12 months, it assesses a recurring phenomenon (6–8).

The significant interaction between waterlogging and chronic illness indicates that chronic illness amplifies the negative association between waterlogging and wellbeing. This pattern is consistent with a syndemic logic, in which environmental stressor interact synergistically rather than additively with health conditions (34,35). While this literature has focused on disease severity as an outcome, our findings extend this synergy to subjective wellbeing, which may feed back into worsening of chronic health conditions (33,34).

Confirming the literature on climate-related gendered vulnerability (35), men reported higher wellbeing than women by about 4 points, as in the EU (36). Yet, the literature points to an underreporting of distress by men (37), suggesting the gender gap may in fact be smaller. Moreover, the waterlogging-gender interaction was not significant (β=-4.86, 95% CI -12.09 to 2.37, SM 5), its direction suggests that men’s wellbeing advantage may narrow with exposure, providing no support for a narrative of amplified female vulnerability to waterlogging. The WHO-5 is more strongly correlated with depressive than anxious symptoms (38); it may be less sensitive to anxiety-related distress, which could differ by gender.

Limitations should be noted. First, our cross-sectional design and 12-month recall window, justified by the drought yet shorter than recall periods used elsewhere in the flood and health literature (26,27), do not allow us to distinguish among the acute, persistent, and anticipatory phases of environmental exposure. Second, non-exposed households may have experienced prior waterlogging, suggesting our estimates may understate the true effect. Third, with only 71 exposed individuals due to the drought, model complexity is constrained and interpretations of non-significant results are limited. Fourth, households were selected based on reporting any symptom in the past month, which may limit representativeness and could correlate with chronic health status.

Finally, as waterlogging intensifies with shrimp farming and climate change, psychosocial support for the vulnerable will be essential, though insufficient without addressing its root causes in land and water management and health inequities. Future work should examine wellbeing as interdependent among household members rather than treating individuals as independent units, alongside the role of social capital in shaping resilience to chronic environmental stress in patrilocal societies.

## Data Availability

All data produced in the present study are available upon reasonable request to the authors

