## Supplementary material for "Waterlogging exposure and mental well-being: a cross-sectional study in rural southwest Bangladesh": SM1

### Supplementary material 1: Sampling and Descriptive statistics

#### Sampling and selection criteria

Households were selected across 10 clusters with a radius from 2 to 3km according to population density, centred on 10 health facilities selected through stratified random sampling from a list of the 53 located in the upazila, to represent the diversity of healthcare levels (primary and secondary), types (private, for-profit, private NGO, public), geographical locations and two levels of exposure to waterlogging (exposed/not exposed)(24). Households were selected if they had 1) any member who had suffered or was suffering from any illness over the last 30 days, or 2) a pregnant woman, or 3) a mother of any child under two years of age. 596 households were selected randomly from a list of 2929 households meeting the inclusion criteria. Within each household, selected participants included, whenever possible, one adult male and one adult female aged 18-59 and 60 years or older. We interviewed a total of 1268 adult individuals from 596 households. Only questionnaires with full information were included (N=1260 respondents, 595 households).

##### Sampling and clusters

Waterlogging exposure was reported in only 3 of 10 clusters (clusters 2, 5, 9), versus 5 expected (SM 3). Despite this small, near-sea-level territory sharing the same (drought-year) rainfall regime, spatial heterogeneity in exposure likely reflects local social-ecological factors. In cluster 9, prior qualitative work (7) identified proximity to shrimp farms, absent or degraded drainage structure, siltation, land-and-water grabbing and heavy rains.

##### Sample characteristics

There is no statistically significant difference in covariate distributions between exposed and non-exposed individuals (Table 1). Mean well-being varies across clusters (SM 3). For the waterlogged clusters, clusters 2 and 9 show among the highest means of wellbeing, while cluster 5 shows the opposite trend. Among clusters with no exposure, cluster 6 stands out as having the lowest means. This heterogeneity confirms the necessity to include the clusters in our models.

Table 1. Sample characteristics by waterlogging exposure status (individual level)

| Terms | Code (Ref cat) | Overall, N = 1260[1] | Exposed household, n = 71[1] | Not exposed household, n = 1189[1] | p-value[2] |
|---|---|---|---|---|---|
| Individual variables | | | | | |
| Wellbeing index (WHO-5) | | 48.5 (20.1) | 46.8 (25.5) | 48.6 (19.8) | 0.559 |
| Wellbeing index ≤28 | | | | | 0.108 |
| ≤28 | 1 | 237 (18.8%) | 19 (26.8%) | 218 (18.3%) | |
| >28 | | 1023 (81.2%) | 52 (73.2%) | 971 (81.7%) | |
| Wellbeing index ≤50 | | | | | 0.689 |
| ≤50 | 1 | 659 (52.3%) | 35 (49.3%) | 624 (52.5%) | |
| >50 | | 601 (47.7%) | 36 (50.7%) | 565 (47.5%) | |
| Age group | | | | | 0.608 |

| | | | | | |
|---|---|---|---|---|---|
| 51+ years old | 1 | 331 (26%) | 21 (30%) | 310 (26%) | |
| 18-50 years old | | 929 (73%) | 50 (70%) | 879 (74%) | |
| Gender | | | | | >0.900 |
| Female | 1 | 648 (51%) | 37 (52%) | 611 (51%) | |
| Male | | 612 (49%) | 34 (48%) | 578 (49%) | |
| Chronic illness status | | | | | 0.437 |
| Chronic illness | 1 | 400 (31.7%) | 26 (36.6%) | 374 (31.5%) | |
| No chronic illness | | 860 (68.3%) | 45 (63.4%) | 815 (68.5%) | |
| Education level | | | | | 0.077[3] |
| No formal education | 1 | 242 (19%) | 16 (23%) | 226 (19%) | |
| Primary | | 378 (30%) | 13 (18%) | 365 (31%) | |
| Secondary | | 594 (47%) | 41 (58%) | 553 (47%) | |
| Higher | | 46 (3.7%) | 1 (1.4%) | 45 (3.8%) | |
| Head of household | | | | | 0.461 |
| Head of household | | 559 (44%) | 28 (39%) | 531 (45%) | |
| Not head | 1 | 701 (56%) | 43 (61%) | 658 (55%) | |
| Microcredit use | | | | | 0.634 |
| User | | 540 (43%) | 28 (39%) | 512 (43%) | |
| Not user | 1 | 720 (57%) | 43(61%) | 677 (57%) | |
| Religion | | | | | 0.150 |
| Hindu | 1 | 312 (25%) | 12 (17%) | 300 (25%) | |
| Muslim | | 948 (75%) | 59 (83%) | 889 (75%) | |
| Household variables | | | | | |
| Household size | | 4.1 (1.4) | 4.2 (1.2) | 4.1 (1.4) | 0.667 |
| Log income | | 9.4 (0.6) | 9.4 (0.6) | 9.4 (0.6) | 0.491 |
| Monthly income (Taka) | | 14089.9 (10365.1) | 15645.1 (12695.1) | 13997.0 (10208.2) | 0.287 |
| Monthly income (std) | | 0 (1) | 0.2 (1.2) | -0.0 (1) | 0.287 |
| Wealth index score (std) | | 0 (1) | 0.2 (1.5) | -0.0 (1) | 0.195 |
| N individuals | 1260 | | | | |
| N households | 595 | | | | |
| N clusters | 10 | | | | |

Notes: [1]Mean (SD); n (%), [2]Welch Two Sample t-test; Pearson's Chi-squared test for the distribution of the variables for exposed/not exposed, [3]Due to low cell counts, Fisher's exact test was used for education level. Voir methodological section for justification of the reference coding.
