## Supplementary material for "Waterlogging exposure and mental well-being: a cross-sectional study in rural southwest Bangladesh": SM2

### Supplementary material 2- alternative models (wealth measures)

Different measures of wealth and transformation are presented in the following table

### Multilevel linear mixed-effects models of wellbeing index (M1 + interaction and alternative models).

Reference category for wealth index: Poorest households.

| | Model of interest M1+ interaction (log income) | | Alternative model with quadratic standardised income | | Alternative model with wealth index quartiles | | Alternative model with wealth index score (standardized) | |
|---|---|---|---|---|---|---|---|---|
| Variable & (reference category) | β_M1i | p_M1i | β_M12 | p_M12 | β_M13 | p_M13 | β_M14 | p_M14 |
| (Intercept) | -18.34 [-40.68, 3.99] | 0.107 | 31.01 [24.67, 37.35] | <0.001 | 26.83 [20.3, 33.36] | <0.001 | 28.4 [22.04, 34.76 | <0.001 |
| Exposure to waterlogging: exposed (not) | -18.31 [-26.45, -10.17] | <0.001 | -18.48 [-26.62, -10.34] | <0.001 | -17.97 [-26.21, -9.73] | <0.001 | -17.9 [-26.14, -9.68] | <0.001 |
| Age group: 18–50 (51+) | 5.02 [2.54, 7.51] | <0.001 | 5.29 [2.8, 7.78] | <0.001 | 5.29 [2.78, 7.79] | <0.001 | 5.21 [2.7, 7.71] | <0.001 |
| Gender: male (female) | 3.95 [0.98, 6.91] | 0.009 | 4.33 [1.37, 7.28] | 0.004 | 4.33 [1.36, 7.3] | 0.004 | 4.49 [1.53, 7.46] | 0.003 |
| Religion: Muslim (Hindu) | 4.77 [1.6, 7.93] | 0.003 | 4.62 [1.45, 7.78] | 0.004 | 4.53 [1.28, 7.77] | 0.006 | 4.31 [1.09, 7.53] | 0.009 |
| Education: primary (No education) | 5.11 [2.12, 8.1] | <0.001 | 5.11 [2.12, 8.1] | <0.001 | 5.14 [2.12, 8.15] | <0.001 | 5.3 [2.29, 8.31] | <0.001 |
| Education: secondary (No education) | 7.7 [4.72, 10.69] | <0.001 | 7.67 [4.68, 10.65] | <0.001 | 7.84 [4.8, 10.88] | <0.001 | 8.21 [5.19, 11.22] | <0.001 |
| Education: higher (No education) | 9.08 [3.21, 14.95] | 0.002 | 8.59 [2.7, 14.48] | 0.004 | 10.16 [4.25, 16.07] | <0.001 | 10.5 [4.64, 16.42] | <0.001 |
| Household monthly income (log) | 5.24 [2.83, 7.65] | <0.001 | | | | | | |

| | Model of interest M1+ interaction (log income) | | Alternative model with quadratic standardised income | | Alternative model with wealth index quartiles | | Alternative model with wealth index score (standardized) | |
|---|---|---|---|---|---|---|---|---|
| Variable & (reference category) | β_M1i | p_M1i | β_M12 | p_M12 | β_M13 | p_M13 | β_M14 | p_M14 |
| Standardised household monthly income | | | 4.44 [2.52, 6.36] | <0.001 | | | | |
| Standardised household monthly income² | | | -0.47 [-0.77, -0.18] | 0.001 | | | | |
| Wealth index score standardised | | | | | | | 0.17 [-1.09, 1.44] | 0.791 |
| Wealth index: Average poor (poorest) | | | | | 2.83 [-0.68, 6.34] | 0.114 | | |
| Wealth index: Average wealthy (poorest) | | | | | 2.69 [-0.87, 6.26] | 0.139 | | |
| Wealth index: Wealthiest (poorest) | | | | | 2.37 [-1.34, 6.08] | 0.211 | | |
| Household size: number of individuals | -0.84 [-1.82, 0.14] | 0.093 | -0.83 [-1.8, 0.14] | 0.093 | -0.09 [-1.02, 0.85] | 0.857 | 0 [-0.93, 0.92] | 0.993 |
| Microcredit: user (not user) | -1.23 [-3.33, 0.87] | 0.252 | -1.08 [-3.19, 1.02] | 0.312 | -1.12 [-3.25, 1.01] | 0.302 | -1.19 [-3.3, 0.93] | 0.273 |
| Household headship: head (not head) | 0.51 [-2.51, 3.52] | 0.742 | 0.18 [-2.83, 3.19] | 0.905 | 0.18 [-2.84, 3.2] | 0.907 | 0.04 [-2.98, 3.06] | 0.978 |
| Chronic illness status: no chronic (chronic illness) | 9.09 [6.87, 11.31] | <0.001 | 9.07 [6.85, 11.29] | <0.001 | 9.09 [6.85, 11.33] | <0.001 | 9 [6.77, 11.24] | <0.001 |
| Waterlogging × No chronic illness | 14.21 [5.48, 22.95] | 0.001 | 14.05 [5.32, 22.79] | 0.002 | 15.31 [6.5, 24.11] | <0.001 | 14.7 [5.94, 23.52] | 0.001 |

| | Model of interest M1+ interaction (log income) | | Alternative model with quadratic standardised income | | Alternative model with wealth index quartiles | | Alternative model with wealth index score (standardized) | |
|---|---|---|---|---|---|---|---|---|
| Variable & (reference category) | β_M1i | p_M1i | β_M12 | p_M12 | β_M13 | p_M13 | β_M14 | p_M14 |
| Cluster 2 | 9.17 [3.69, 14.65] | 0.001 | 9.64 [4.11, 15.16] | <0.001 | 8.08 [2.52, 13.65] | 0.004 | 8.32 [2.76, 13.88] | 0.003 |
| Cluster 3 | 0.69 [-4.6, 5.99] | 0.797 | 0.73 [-4.56, 6.02] | 0.786 | -0.34 [-5.7, 5.02] | 0.901 | -0.4 [-5.76, 4.96] | 0.884 |
| Cluster 4 | -0.94 [-6.28, 4.4] | 0.730 | -0.91 [-6.24, 4.42] | 0.738 | -2.51 [-7.88, 2.87] | 0.361 | -2.45 [-7.84, 2.93] | 0.372 |
| Cluster 5 | -5.02 [-10.52, 0.48] | 0.073 | -5.07 [-10.56, 0.42] | 0.070 | -6.79 [-12.38, -1.2] | 0.017 | -6.44 [-12, -0.89] | 0.023 |
| Cluster 6 | -6.42 [-11.87, -0.96] | 0.021 | -6.14 [-11.6, -0.68] | 0.027 | -8.74 [-14.24, -3.25] | 0.002 | -8.29 [-13.76, -2.81] | 0.003 |
| Cluster 7 | 2.73 [-2.74, 8.2] | 0.328 | 2.83 [-2.64, 8.29] | 0.310 | 1.41 [-4.14, 6.97] | 0.618 | 1.74 [-3.82, 7.3] | 0.539 |
| Cluster 8 | 0.92 [-4.33, 6.17] | 0.731 | 0.98 [-4.26, 6.22] | 0.714 | -0.79 [-6.12, 4.54] | 0.772 | -0.45 [-5.77, 4.88] | 0.869 |
| Cluster 9 | 10.55 [4.78, 16.33] | <0.001 | 10.54 [4.78, 16.3] | <0.001 | 7.96 [2.2, 13.71] | 0.007 | 7.99 [2.24, 13.74] | 0.007 |
| Cluster 10 | -0.08 [-5.37, 5.22] | 0.977 | -0.03 [-5.32, 5.25] | 0.990 | -1.26 [-6.67, 4.15] | 0.647 | -1.09 [-6.47, 4.3] | 0.692 |
| *Number of observations* | *1260* | | *1260* | | *1260* | | *1260* | |
| *Number of groups* | *595* | | *595* | | *595* | | *595* | |
| *Random-effect variance* | *104.15* | | *103.5* | | *110.4* | | *111.1* | |
| *Residual variance* | *218.51* | | *218.59* | | *218.5* | | *218.3* | |

| | Model of interest M1+ interaction (log income) | | Alternative model with quadratic standardised income | | Alternative model with wealth index quartiles | | Alternative model with wealth index score (standardized) | |
|---|---|---|---|---|---|---|---|---|
| Variable & (reference category) | β_M1i | p_M1i | β_M12 | p_M12 | β_M13 | p_M13 | β_M14 | p_M14 |
| *ICC* | *0.323* | | *0.321* | | *0.336* | | *0.337* | |
| *Marginal $R^2$* | *0.208* | | *0.21* | | *0.192* | | *0.190* | |
| *Conditional $R^2$* | *0.464* | | *0.464* | | *0.463* | | *0.463* | |
| *AIC* | *10736.1* | | *10738.9* | | *10748.8* | | *10755.3* | |

**Comparison of models for different measures of wealth**

Economic status may influence perceptions of wellbeing, and can be measured in several ways. In rural Bangladesh, where income may be seasonal, irregular and vulnerable to environmental shocks, both dimensions are relevant. Current income may reflect immediate difficulties in livelihood security, while asset ownership captures accumulated wealth over time, which may buffer against short-term financial strain. Our model of interest, uses a log transformation ((log(income + 1)) of monthly household income, given its right-skewed distribution. To compare with our log model of interest (Model 1 with interactions, AIC = 10736.08, df = 25), we examined the potential non-linear association between income and wellbeing in an alternative model using a quadratic term of standardised monthly income ( model with quadratic term, AIC= 10738.95, df=26). Both models were compared using the Akaike Information Criterion. Adding a quadratic term did not substantially improve the models (likelihood ratio, chi2(1)=2.470, p=0.116, ΔAIC=1).

Due to the low and middle-income countries and rural context, two additional alternative models with asset-based measures of socioeconomic status instead of household income were also estimated: one using quartiles of wealth index (based on DHS wealth index construction, see descriptive statistics below) (AIC = 10748.84, df = 27) and one using the continuous standardised wealth index score (AIC= 10755.26, df=25). Both models showed poorer fit compared to the log-income model (ΔAIC = 12.76 and 19.18, respectively). These results confirm that the log-transformed income model is the best-fitting model for our data. The log-transformed choice is also supported by the literature on income and subjective wellbeing in rural Bangladesh (Tauseef 2022).

### Descriptive statistics for wealth categories

| Terms | Code (Ref cat) | Overall, N = 1260[1] | Exposed, n = 71[1] | Not exposed, n = 1189[1] | p-value[2] |
|---|---|---|---|---|---|
| Household variables | | | | | |
| Log income | | 9.4 (0.6) | 9.4 (0.6) | 9.4 (0.6) | 0.491 |
| Monthly income (Taka) | | 14089.9 (10365.1) | 15645.1 (12695.1) | 13997.0 (10208.2) | 0.287 |
| Monthly income (std) | | 0 (1) | 0.2 (1.2) | -0.0 (1) | 0.287 |
| Wealth index score (std) | | 0 (1) | 0.2 (1.5) | -0.0 (1) | 0.195 |
| Wealth index quartiles | | | | | 0.0004[3] |
| Poorest | 1 | (22.5%) | 23 (32.4%) | 261 (21.9%) | |
| Average poor | | (25%) | 5 (7%) | 310 (26.1%) | |
| Average wealthy | | (25.1%) | 24 (33.8%) | 292 (24.6%) | |
| Wealthiest | | (27.4%) | 19 (26.8%) | 326 (27.4%) | |
| N individuals | 1260 | | | | |
| N households | 595 | | | | |
| N clusters | 10 | | | | |

Notes: [1]Mean (SD); n (%), [2]Welch Two Sample t-test; Pearson's Chi-squared test for the distribution of the variables for exposed/not exposed, [3]Due to low cell counts, Fisher’s exact test was used for education level. Voir methodological section for justification of the reference coding.
