## Supplementary material for "Waterlogging exposure and mental well-being: a cross-sectional study in rural southwest Bangladesh": SM3

### Supplementary material 3: Information on clusters

Sampling was organised in 10 clusters (see SM1). Below is information about these 10 clusters

Waterlogging exposure by cluster, planned design vs reported exposure

| Cluster | Planned design | N total individuals (HH) | Number of not exposed individuals (HH) | Number of exposed individuals (HH) | % exposed individuals (%HH) |
|---|---|---|---|---|---|
| 1 | Not waterlogged | 127 (59) | 127 (59) | 0 (0) | 0% |
| 2 | Waterlogged | 125 (59) | 98 (46) | 27 (13) | 21.6% (22%) |
| 3 | Not waterlogged | 129 (60) | 129 (60) | 0 (0) | 0% |
| 4 | Waterlogged | 126 (60) | 126 (60) | 0 (0) | 0% |
| 5 | Waterlogged | 119 (58) | 113 (56) | 6 (2) | 5% (3.4%) |
| 6 | Not waterlogged | 121 (60) | 121 (60) | 0 (0) | 0% |
| 7 | Not waterlog)ged | 117 (59) | 117 (59) | 0 (0) | 0% |
| 8 | Not waterlogged | 141 (60) | 141 (60) | 0 (0) | 0% |
| 9 | Waterlogged | 125 (60) | 87 (42) | 38 (18) | 30.4% (30%) |
| 10 | Waterlogged | 130 (60) | 130 (60) | 0 (0) | 0% |
| N total 10 | | 1260 (595) | 1189 (562) | 71 (33) | 5.6% (5.5%) |

*Notes: HH = number of households. Grey shading = clusters with reported waterlogging exposure, included in robustness analyses (M2). All 7 other clusters were included in robustness analysis (M3)*

##### Wealth index quartile distribution by cluster (%)

| Wealth index quartile | Cluster 1 | Cluster 2 | Cluster 3 | Cluster 4 | Cluster 5 | Cluster 6 | Cluster 7 | Cluster 8 | Cluster 9 | Cluster 10 |
|---|---|---|---|---|---|---|---|---|---|---|
| Poorest | 27.6 | 18.4 | 33.3 | 28.6 | 17.6 | 14 | 20.5 | 9.2 | 38.4 | 18.5 |
| Average_Poor | 26 | 31.2 | 35.7 | 28.6 | 16.8 | 33.1 | 30.8 | 19.9 | 17.6 | 11.5 |
| Average_Wealthy | 21.3 | 15.2 | 14.7 | 21.4 | 33.6 | 25.6 | 23.9 | 31.2 | 34.4 | 29.2 |
| Wealthiest | 25.2 | 35.2 | 16.3 | 21.4 | 31.9 | 27.3 | 24.8 | 39.7 | 9.6 | 40.8 |
| **Waterlogging exposure** | **No** | **Yes** | **No** | **No** | **Yes** | **No** | **No** | **No** | **Yes** | **No** |

Figure : Mean well-being score varies across clusters, indicating differences between clusters. Mean +95%CI

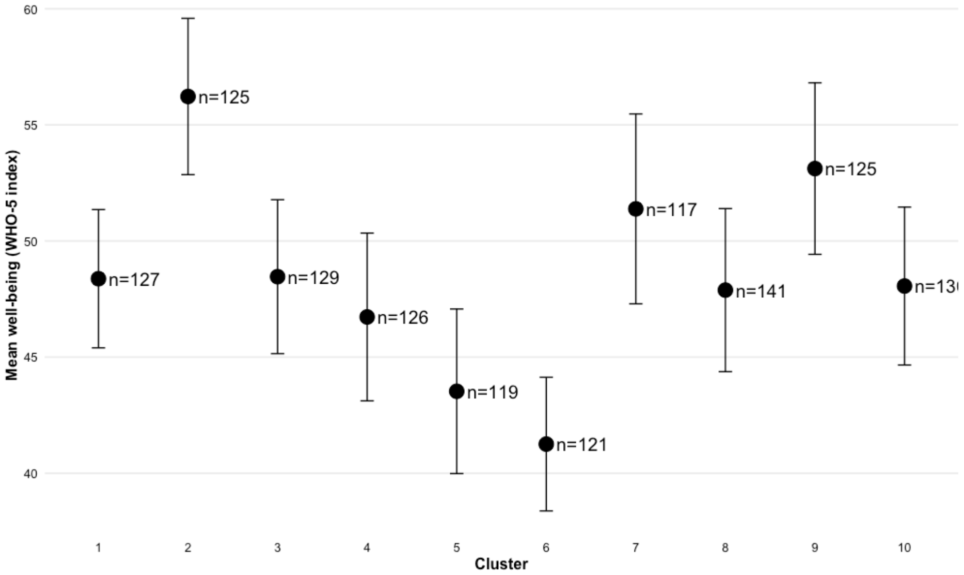


**Marginal means derived from model 1 with interaction.**

The ten clusters correspond to our selected sub-samples. Living in certain clusters is significantly associated with higher or lower well-being (SM 3). In Figure 2, marginal means, derived from the regression (Model 1 + interactions; Table 1), indicate that these differences persist even after accounting for individual and household characteristics. This suggests the importance of contextual, cluster-level factors (such as infrastructure, past waterlogging exposure, market integration, power dynamics, social capital etc.) that are not captured in these analyses.

**Figure 2: Marginal means (+IC 95%) derived from model 1 with interaction.**

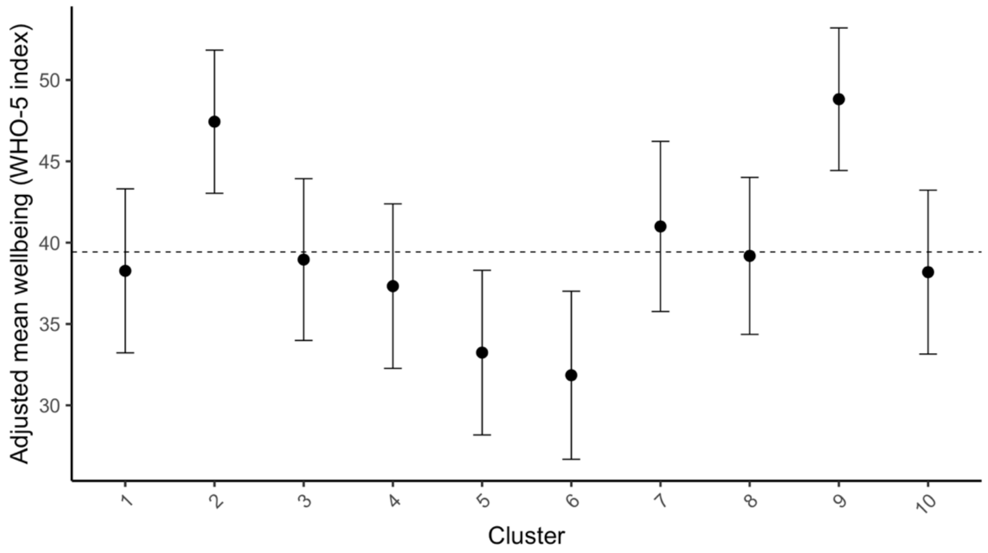

Local context

Visual inspection of satellite images (Figure 1A and 1B) taken during the data collection (8 September 2022) highlights the proximity to shrimp farming enclosures (in black in Figure 1B), and selected households from clusters 2, 5 and 9 seem to be surrounded by enclosures, which might explain why, even despite the drought, they reported waterlogging in the past 12 months. However, other characteristics should also be considered as explained earlier (drainage, degradation/siltation of the canals, type and density of enclosures etc).

Figures 1 A and 1B. Cluster and waterlogging exposure at the household level, in Tala upazila

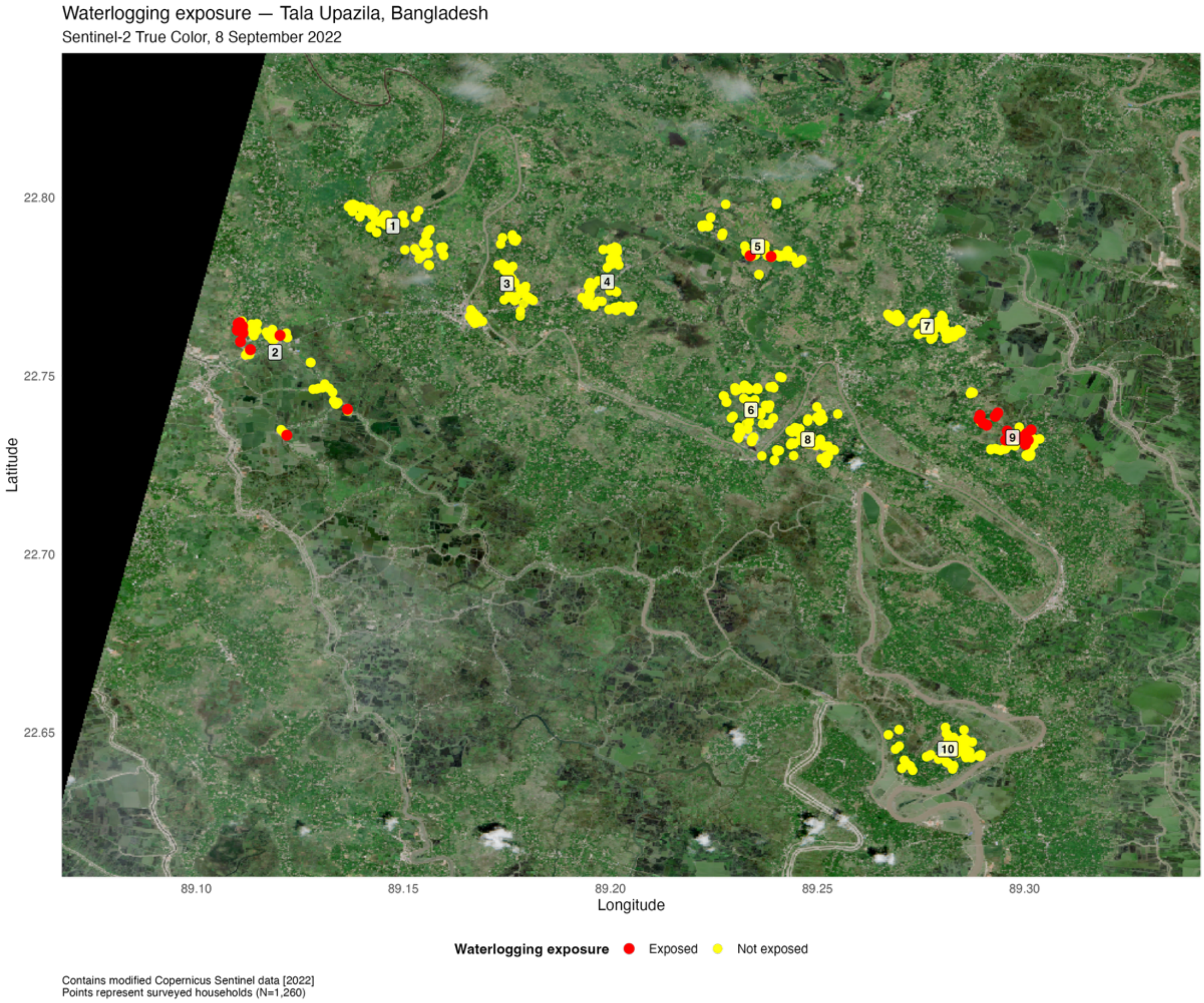

Waterlogging exposure — Tala Upazila, Bangladesh

Sentinel-2 False Color, 8 September 2022

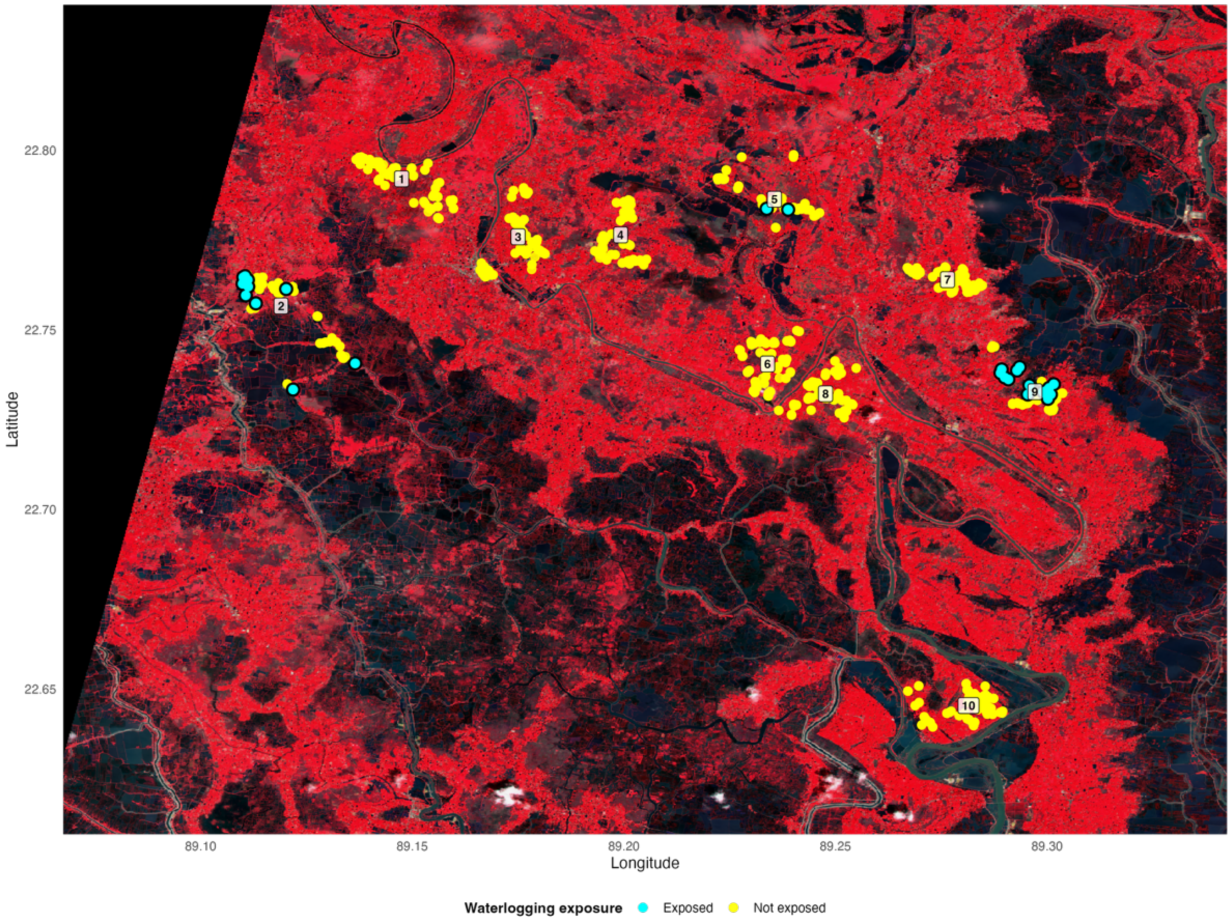


Contains modified Copernicus Sentinel data [2022]
Points represent surveyed households (N=1,260)
False color: vegetation = red, water/aquaculture = blue/black
