## Supplementary material for "Waterlogging exposure and mental well-being: a cross-sectional study in rural southwest Bangladesh": SM4

### Supplementary material 4: Multilevel linear mixed-effects models of wellbeing index (clusters as fixed or random effects)

Reference category for wealth index: Poorest households.

| | Model of interest M1+ interaction | | Alternative model | |
|---|---|---|---|---|
| | **Clusters as fixed effects** | | **Clusters as random effects** | |
| Variable & (reference category) | β_M1i | p_M1i | β_M14 | p_M14 |
| (Intercept) | -18.34 [-40.68, 3.99] | 0.107 | -16.51 [-38.18, 5.15] | 0.135 |
| Exposure to waterlogging: exposed (not) | -18.31 [-26.45, -10.17] | <0.001 | -16.84 [-24.9, -8.79] | <0.001 |
| Age group: 18–50 (51+) | 5.02 [2.54, 7.51] | <0.001 | 5.03 [2.54, 7.51] | <0.001 |
| Gender: male (female) | 3.95 [0.98, 6.91] | 0.009 | 3.98 [1.02, 6.95] | 0.009 |
| Religion: Muslim (Hindu) | 4.77 [1.6, 7.93] | 0.003 | 4.44 [1.32, 7.55] | 0.005 |
| Education: primary (No education) | 5.11 [2.12, 8.1] | <0.001 | 4.99 [2, 7.97] | 0.001 |
| Education: secondary (No education) | 7.7 [4.72, 10.69] | <0.001 | 7.55 [4.57, 10.54] | <0.001 |
| Education: higher (No education) | 9.08 [3.21, 14.95] | 0.002 | 8.84 [2.97, 14.7] | 0.003 |
| Household monthly income (log) | 5.24 [2.83, 7.65] | <0.001 | 5.2 [2.8, 7.59] | <0.001 |
| Household size: number of individuals | -0.84 [-1.82, 0.14] | 0.093 | -0.86 [-1.84, 0.12] | 0.086 |
| Microcredit: user (not user) | -1.23 [-3.33, 0.87] | 0.252 | -1.22 [-3.32, 0.88] | 0.254 |
| Household headship: head (not head) | 0.51 [-2.51, 3.52] | 0.742 | 0.45 [-2.56, 3.47] | 0.769 |

| | Model of interest<br>M1+ interaction | | Alternative model | |
|---|---|---|---|---|
| | **Clusters as fixed effects** | | **Clusters as random effects** | |
| Variable & (reference category) | β_M1i | p_M1i | β_M14 | p_M14 |
| Chronic illness status: no chronic (chronic illness) | 9.09 [6.87, 11.31] | <0.001 | 9.23 [7.01, 11.45] | <0.001 |
| Waterlogging × No chronic illness | 14.21 [5.48, 22.95] | 0.001 | 14.07 [5.34, 22.8] | 0.002 |
| Cluster 2 | 9.17 [3.69, 14.65] | 0.001 | | |
| Cluster 3 | 0.69 [-4.6, 5.99] | 0.797 | | |
| Cluster 4 | -0.94 [-6.28, 4.4] | 0.730 | | |
| Cluster 5 | -5.02 [-10.52, 0.48] | 0.073 | | |
| Cluster 6 | -6.42 [-11.87, -0.96] | 0.021 | | |
| Cluster 7 | 2.73 [-2.74, 8.2] | 0.328 | | |
| Cluster 8 | 0.92 [-4.33, 6.17] | 0.731 | | |
| Cluster 9 | 10.55 [4.78, 16.33] | <0.001 | | |
| Cluster 10 | -0.08 [-5.37, 5.22] | 0.977 | | |
| *Number of observations* | *1260* | | *1260* | |
| *Number of groups* | *595* | | *595* | |
| *Random-effect variance* | *104.15* | | *104* | |
| *Residual variance* | *218.51* | | *218.64* | |
| *ICC* | *0.323* | | *0.369* | |

| | Model of interest<br>M1+ interaction | | Alternative model | |
|---|---|---|---|---|
| | **Clusters as fixed effects** | | **Clusters as random effects** | |
| Variable & (reference category) | β_M1i | p_M1i | β_M14 | p_M14 |
| *Marginal R²* | *0.208* | | *0.165* | |
| *Conditional R²* | *0.464* | | *0.473* | |
| *AIC* | *10736.1* | | *10777.9* | |

**Notes:** Clusters were purposively selected to balance waterlogging exposed and non-exposed areas. Only three of ten (instead of five) included households reporting waterlogging in the past year, likely reflecting drought conditions at the time of data collection. This still supports two analytical contrasts: between exposed and non-exposed clusters (M2 and M4, table 1 in the manuscript), and, within exposed clusters, between households that report exposure and those that do not (model M2, table 1 in the manuscript).
Given the limited number of clusters (10), estimation of cluster-level random effects may be unreliable. Clusters may represent different communities selected around different categories of health structures. While the type of health structures might not be a directly important criterion for mental wellbeing, clusters may nevertheless reflect different levels of cluster development or/and local social context (local power dynamics, support network), that may be important for wellbeing.
When both models are performed, results appear consistent across models, but the model with fixed clusters presents a better fit compared to the model with random effects (AIC M1i fixed: 10736.08, AIC M1i random: 10777.83, ΔAIC=41.9). A likelihood ratio test confirmed that fixed effects clusters present a significantly better fit than random effects clusters ($\chi^2$ = 29.69, df=8, $p < 0.001$, ΔAIC=13).
