## Supplementary material for "Waterlogging exposure and mental well-being: a cross-sectional study in rural southwest Bangladesh": SM5

#### Supplementary material 5 – Model 2 -only clusters with reported waterlogging

### Table. Multilevel linear mixed-effects models of WHO-5 well-being index. Sensitivity analyses for Model 2.

Notes: coefficients are presented with 95% confidence intervals.

| | M2 with 3 clusters (2, 5, 9) | | M2 with 2 clusters (2, 9) | |
|---|---|---|---|---|
| Variable | β_M2_3clusters | p_M2_3clusters | β_M2_2clusters | p_M2_2clusters |
| (Intercept) | 4.67 [-31.37, 40.72] | 0.798 | 16.29 [-27.06, 59.63] | 0.458 |
| Exposure to waterlogging: exposed (ref: non-exposed) | -17.25 [-26.19, -8.32] | <0.001 | -18.1 [-28.29, -7.91] | <0.001 |
| Age group: 18–50 (ref: 51+) | 5.67 [1.2, 10.14] | 0.013 | 6.07 [0.46, 11.68] | 0.034 |
| Gender: Male (ref: female) | 3.14 [-3.02, 9.29] | 0.316 | 2.99 [-4.84, 10.81] | 0.453 |
| Religion: Muslim (ref: Hindu) | 1.72 [-4.97, 8.4] | 0.613 | 3.65 [-3.19, 10.49] | 0.292 |
| Education: Primary (ref: no formal education) | 4.46 [-0.7, 9.62] | 0.090 | 4.32 [-2.21, 10.84] | 0.194 |
| Education: Secondary (ref: no formal education) | 7.9 [2.74, 13.06] | 0.003 | 7.55 [1.04, 14.06] | 0.023 |
| Education: Higher (ref: no formal education) | 6.45 [-4.47, 17.37] | 0.246 | 12.14 [-7.55, 31.84] | 0.226 |
| Household monthly income (log) | 3.79 [-0.17, 7.74] | 0.060 | 3.05 [-1.72, 7.83] | 0.207 |
| Household size: number of individuals | -0.86 [-2.76, 1.04] | 0.375 | -1.67 [-4.08, 0.75] | 0.174 |
| Microcredit use: User (ref: non-user) | 0.71 [-3.07, 4.49] | 0.711 | -0.36 [-5.15, 4.44] | 0.884 |
| Household headship: Head (ref: non-head) | 2.74 [-3.56, 9.05] | 0.392 | 2.61 [-5.37, 10.59] | 0.520 |

| | M2 with 3 clusters (2, 5, 9) | | M2 with 2 clusters (2, 9) | |
|---|---|---|---|---|
| Variable | β_M2_3clusters | p_M2_3clusters | β_M2_2clusters | p_M2_2clusters |
| Chronic illness status: No chronic illness (ref: chronic illness) | 8.72 [3.72, 13.72] | <0.001 | 6.65 [-0.16, 13.47] | 0.055 |
| Waterlogging × No chronic illness | 13.45 [3.88, 23.01] | 0.006 | 13.37 [2.17, 24.58] | 0.020 |
| Cluster 5 (ref: cluster 2) | -13.02 [-19.59, -6.46] | <0.001 | | |
| Cluster 9 (ref: cluster 2) | 1.86 [-4.46, 8.17] | 0.562 | 0.34 [-6.08, 6.75] | 0.917 |
| *Number of observations* | *369* | | *250* | |
| *Number of groups* | *177* | | *119* | |
| *Random-effect variance* | *122.48* | | *105.14* | |
| *Residual variance* | *188.61* | | *211.7* | |
| *ICC* | *0.394* | | *0.332* | |
| *Marginal $R^2$* | *0.279* | | *0.229* | |
| *Conditional $R^2$* | *0.563* | | *0.485* | |
| *AIC* | *3094.1* | | *2092.7* | |
