## Supplementary material for "Waterlogging exposure and mental well-being: a cross-sectional study in rural southwest Bangladesh": SM6

### Supplementary material 6: Alternative models with Interactions — WHO-5 wellbeing index

| Model | MA1 | | MA2 | | MA3 | | MA4 | |
|---|---|---|---|---|---|---|---|---|
| **Interaction of interest: waterlogging and** | **Age** | | **Gender** | | **Education** | | **Income** | |
| Variable | β | p | β | p | β | p | β | p |
| (Intercept) | -18.46 [-40.82, 3.89] | 0.105 | -18.54 [-40.86, 3.79] | 0.104 | -18.63 [-40.9, 3.64] | 0.101 | -17.56 [-40.56, 5.44] | 0.134 |
| Exposure to waterlogging: exposed (not) | -21.61 [-31.24, -11.99] | <0.001 | -16.16 [-24.9, -7.41] | <0.001 | -21.01 [-31.59, -10.43] | <0.001 | -29.32 [-106.09, 47.44] | 0.453 |
| Age group: 18–50 (51+) | 4.73 [2.2, 7.26] | <0.001 | 5.03 [2.54, 7.52] | <0.001 | 5.06 [2.57, 7.54] | <0.001 | 5.03 [2.54, 7.52] | <0.001 |
| Gender: male (female) | 3.84 [0.87, 6.81] | 0.011 | 4.23 [1.23, 7.22] | 0.006 | 4.03 [1.08, 6.98] | 0.008 | 3.95 [0.98, 6.92] | 0.009 |
| Religion: Muslim (Hindu) | 4.75 [1.58, 7.93] | 0.003 | 4.77 [1.6, 7.94] | 0.003 | 4.74 [1.57, 7.91] | 0.003 | 4.77 [1.6, 7.94] | 0.003 |
| Education: primary (no education) | 5.08 [2.09, 8.06] | <0.001 | 5.03 [2.05, 8.02] | <0.001 | | | 5.11 [2.12, 8.09] | <0.001 |
| Education: secondary (no education) | 7.65 [4.66, 10.64] | <0.001 | 7.65 [4.66, 10.63] | <0.001 | | | 7.71 [4.72, 10.7] | <0.001 |
| Education: higher (no education) | 8.99 [3.12, 14.86] | 0.003 | 9.02 [3.15, 14.89] | 0.003 | | | 9.1 [3.23, 14.97] | 0.002 |
| Education: primary (no education) | | | | | 4.82 [1.76, 7.89] | 0.002 | | |
| Education: secondary or higher (no education) | | | | | 7.53 [4.48, 10.58] | <0.001 | | |
| Household monthly income (log) | 5.27 [2.86, 7.68] | <0.001 | 5.25 [2.85, 7.66] | <0.001 | 5.29 [2.89, 7.69] | <0.001 | 5.16 [2.68, 7.63] | <0.001 |

| Model | MA1 | | MA2 | | MA3 | | MA4 | |
|---|---|---|---|---|---|---|---|---|
| **Interaction of interest: waterlogging and** | **Age** | | **Gender** | | **Education** | | **Income** | |
| Variable | β | p | β | p | β | p | β | p |
| Household size (number of individuals) | -0.82 [-1.8, 0.17] | 0.103 | -0.84 [-1.82, 0.14] | 0.093 | -0.85 [-1.83, 0.13] | 0.089 | -0.84 [-1.82, 0.14] | 0.094 |
| Microcredit: user (not user) | -1.31 [-3.42, 0.79] | 0.221 | -1.29 [-3.4, 0.81] | 0.229 | -1.26 [-3.36, 0.85] | 0.242 | -1.24 [-3.34, 0.87] | 0.249 |
| Household headship: head (not head) | 0.66 [-2.36, 3.69] | 0.666 | 0.51 [-2.51, 3.52] | 0.741 | 0.5 [-2.51, 3.52] | 0.743 | 0.5 [-2.51, 3.52] | 0.744 |
| Chronic illness status: no chronic (chronic) | 9.16 [6.93, 11.38] | <0.001 | 9.07 [6.84, 11.29] | <0.001 | 9.1 [6.88, 11.33] | <0.001 | 9.09 [6.86, 11.31] | <0.001 |
| Waterlogging × No chronic illness | 12.29 [3.07, 21.51] | 0.009 | 14.43 [5.69, 23.17] | 0.001 | 13.34 [4.37, 22.32] | 0.004 | 13.95 [5.04, 22.86] | 0.002 |
| Waterlogging × Age (18–50) | 6.27 [-3.45, 16] | 0.206 | | | | | | |
| Waterlogging × Gender (male) | | | -4.86 [-12.09, 2.37] | 0.188 | | | | |
| Waterlogging × Education (primary) | | | | | 4.86 [-8.07, 17.8] | 0.461 | | |
| Waterlogging × Education (secondary or higher) | | | | | 3.92 [-6.9, 14.75] | 0.477 | | |
| Waterlogging × Household income (log) | | | | | | | 1.19 [-7.03, 9.4] | 0.777 |
| Cluster 2 | 9.08 [3.59, 14.56] | 0.001 | 9.21 [3.73, 14.68] | 0.001 | 9.16 [3.67, 14.64] | 0.001 | 9.14 [3.65, 14.62] | 0.001 |
| Cluster 3 | 0.7 [-4.6, 6] | 0.796 | 0.69 [-4.6, 5.98] | 0.799 | 0.73 [-4.56, 6.02] | 0.786 | 0.68 [-4.62, 5.98] | 0.802 |

| Model | MA1 | | MA2 | | MA3 | | MA4 | |
|---|---|---|---|---|---|---|---|---|
| **Interaction of interest: waterlogging and** | **Age** | | **Gender** | | **Education** | | **Income** | |
| Variable | β | p | β | p | β | p | β | p |
| Cluster 4 | -0.94 [-6.28, 4.41] | 0.731 | -0.94 [-6.28, 4.4] | 0.728 | -0.92 [-6.26, 4.43] | 0.736 | -0.97 [-6.32, 4.39] | 0.723 |
| Cluster 5 | -5.03 [-10.54, 0.47] | 0.073 | -5.02 [-10.52, 0.48] | 0.074 | -4.97 [-10.46, 0.52] | 0.076 | -5.11 [-10.65, 0.43] | 0.070 |
| Cluster 6 | -6.42 [-11.87, -0.96] | 0.021 | -6.42 [-11.87, -0.97] | 0.021 | -6.37 [-11.82, -0.91] | 0.022 | -6.45 [-11.92, -0.99] | 0.021 |
| Cluster 7 | 2.72 [-2.75, 8.2] | 0.329 | 2.72 [-2.75, 8.19] | 0.330 | 2.77 [-2.7, 8.24] | 0.320 | 2.7 [-2.78, 8.19] | 0.333 |
| Cluster 8 | 0.9 [-4.36, 6.16] | 0.736 | 0.92 [-4.33, 6.17] | 0.730 | 0.95 [-4.31, 6.2] | 0.724 | 0.9 [-4.36, 6.16] | 0.736 |
| Cluster 9 | 10.6 [4.82, 16.38] | <0.001 | 10.5 [4.72, 16.27] | <0.001 | 10.57 [4.79, 16.35] | <0.001 | 10.59 [4.8, 16.38] | <0.001 |
| Cluster 10 | -0.1 [-5.4, 5.2] | 0.971 | -0.09 [-5.38, 5.2] | 0.973 | -0.06 [-5.36, 5.24] | 0.982 | -0.1 [-5.4, 5.2] | 0.970 |
| *Number of observations* | *1260* | | *1260* | | *1260* | | *1260* | |
| *Number of households* | *595* | | *595* | | *595* | | *595* | |
| *Residual variance* | *218.01* | | *218.43* | | *218.52* | | *218.46* | |
| *ICC (household level)* | *0.324* | | *0.323* | | *0.323* | | *0.324* | |
| *Marginal $R^2$* | *0.209* | | *0.209* | | *0.208* | | *0.208* | |
| *Conditional $R^2$* | *0.466* | | *0.464* | | *0.464* | | *0.464* | |
| *AIC* | *10731.4* | | *10731.9* | | *10731.1* | | *10733.3* | |

Note: For MA3, secondary level and higher education were merged, due to low count of waterlogged*higher education, to allow for testing the interaction waterlogging*education (variable with 3 categories instead of 4 categories)
