## Supplementary material for "Waterlogging exposure and mental well-being: a cross-sectional study in rural southwest Bangladesh": SM7

### Supplementary material 7. Generalised linear mixed logistic regression models — WHO-5 well-being index

| | WHO-5 ≤50 (with interaction) | | WHO-5 ≤50 (without interaction) | | WHO-5 ≤28 (without interaction) | |
|---|---|---|---|---|---|---|
| Variable | OR (95% CI) | p-value | OR (95% CI) | p-value | OR (95% CI) | p-value |
| (Intercept) | 11.55 [4.3, 31] | <0.001 | 12.08 [4.48, 32.56] | <0.001 | 1.21 [0.39, 3.81] | 0.742 |
| Exposure to waterlogging: exposed (not) | 5.4 [1.37, 21.36] | 0.016 | 2.43 [1.03, 5.69] | 0.042 | 7.25 [2.31, 22.79] | <0.001 |
| Age group: 18–50 (51+) | 0.53 [0.35, 0.79] | 0.002 | 0.52 [0.35, 0.78] | 0.002 | 0.41 [0.25, 0.67] | <0.001 |
| Gender: male (female) | 0.56 [0.34, 0.92] | 0.022 | 0.57 [0.35, 0.93] | 0.024 | 0.66 [0.36, 1.19] | 0.170 |
| Religion: Muslim (Hindu) | 0.39 [0.24, 0.62] | <0.001 | 0.39 [0.24, 0.63] | <0.001 | 0.68 [0.38, 1.22] | 0.195 |
| Education: primary (No education) | 0.57 [0.35, 0.93] | 0.024 | 0.57 [0.35, 0.92] | 0.023 | 0.57 [0.32, 0.99] | 0.047 |
| Education: secondary (No education) | 0.34 [0.21, 0.55] | <0.001 | 0.34 [0.2, 0.55] | <0.001 | 0.37 [0.21, 0.67] | <0.001 |
| Education: higher (No education) | 0.5 [0.2, 1.28] | 0.148 | 0.51 [0.2, 1.3] | 0.160 | 0.6 [0.18, 1.95] | 0.393 |
| Household monthly income (standardised) | 0.5 [0.37, 0.67] | <0.001 | 0.49 [0.36, 0.66] | <0.001 | 0.68 [0.43, 1.09] | 0.111 |
| Household monthly income$^2$ (standardised) | 1.07 [1.03, 1.12] | <0.001 | 1.08 [1.03, 1.12] | <0.001 | 0.96 [0.8, 1.16] | 0.695 |
| Household size: number of individuals | 1.15 [1, 1.33] | 0.054 | 1.15 [1, 1.33] | 0.051 | 1.13 [0.94, 1.35] | 0.191 |

| | WHO-5 ≤50 (with interaction) | | WHO-5 ≤50 (without interaction) | | WHO-5 ≤28 (without interaction) | |
|---|---|---|---|---|---|---|
| Variable | OR (95% CI) | p-value | OR (95% CI) | p-value | OR (95% CI) | p-value |
| Microcredit user (not user) | 1.4 [1.01, 1.94] | 0.046 | 1.41 [1.01, 1.96] | 0.042 | 0.94 [0.62, 1.44] | 0.790 |
| Household headship: head (not head) | 0.95 [0.58, 1.56] | 0.834 | 0.94 [0.57, 1.54] | 0.801 | 0.73 [0.4, 1.34] | 0.313 |
| Chronic illness status: no chronic illness (chronic illness) | 0.33 [0.23, 0.48] | <0.001 | 0.31 [0.21, 0.44] | <0.001 | 0.27 [0.17, 0.41] | <0.001 |
| Waterlogging × No chronic illness | 0.31 [0.07, 1.41] | 0.129 | | | | |
| Cluster 2 | 0.28 [0.12, 0.63] | 0.002 | 0.27 [0.12, 0.62] | 0.002 | 0.23 [0.07, 0.76] | 0.016 |
| Cluster 3 | 0.92 [0.43, 1.98] | 0.838 | 0.92 [0.43, 1.98] | 0.830 | 0.71 [0.26, 1.93] | 0.505 |
| Cluster 4 | 1.03 [0.48, 2.22] | 0.945 | 1.02 [0.47, 2.2] | 0.968 | 1.54 [0.59, 4] | 0.380 |
| Cluster 5 | 1.99 [0.89, 4.45] | 0.092 | 2 [0.89, 4.48] | 0.093 | 1.82 [0.68, 4.89] | 0.234 |
| Cluster 6 | 2.57 [1.13, 5.84] | 0.024 | 2.56 [1.12, 5.83] | 0.026 | 1.39 [0.52, 3.72] | 0.514 |
| Cluster 7 | 0.59 [0.27, 1.31] | 0.196 | 0.59 [0.26, 1.3] | 0.189 | 1.61 [0.6, 4.29] | 0.343 |
| Cluster 8 | 0.76 [0.36, 1.6] | 0.465 | 0.76 [0.35, 1.62] | 0.472 | 0.89 [0.35, 2.27] | 0.800 |
| Cluster 9 | 0.27 [0.11, 0.62] | 0.002 | 0.27 [0.12, 0.63] | 0.003 | 0.18 [0.05, 0.65] | 0.008 |
| Cluster 10 | 0.98 [0.46, 2.1] | 0.961 | 0.97 [0.45, 2.09] | 0.942 | 0.84 [0.32, 2.22] | 0.724 |
| *Number of observations* | *1260* | | *1260* | | *1260* | |
| *Number of households* | *595* | | *595* | | *595* | |
| *Household-level variance* | *1.4* | | *1.42* | | *1.85* | |

| | WHO-5 ≤50 (with interaction) | WHO-5 ≤50 (without interaction) | | | WHO-5 ≤28 (without interaction) | |
|---|---|---|---|---|---|---|
| Variable | OR (95% CI) | p-value | OR (95% CI) | p-value | OR (95% CI) | p-value |
| *AIC* | *1536.1* | | *1536.5* | | *1076* | |

OR: Odds Ratio; CI: 95% Confidence Interval. OR < 1 indicates lower odds of poor wellbeing relative to the reference category, whereas OR > 1 indicates higher odds of poor wellbeing.
WHO-5 ≤50: validated clinical threshold for poor well-being.
WHO-5 ≤28: threshold for severe poor well-being.
Models estimated using generalised linear mixed-effects logistic regression (glmer, bobyqa optimiser, lme4 package, R version 4.3.0).
Reference categories: no waterlogging exposure, age ≥51 years, female, Hindu religion, no formal education, not microcredit user, not head of household, chronic illness present.
Cluster 1 as reference for cluster fixed effects.
